# Inference of core needle biopsy whole slide images requiring definitive therapy for prostate cancer

**DOI:** 10.1101/2022.09.06.22279630

**Authors:** Masayuki Tsuneki, Makoto Abe, Shin Ichihara, Fahdi Kanavati

## Abstract

Prostate cancer is often a slowly progressive indolent disease. Unnecessary treatments from overdiagnosis are a significant concern, particularly low-grade disease. Active surveillance has being considered as a risk management strategy to avoid potential side effects by unnecessary radical treatment. In 2016, American Society of Clinical Oncology (ASCO) endorsed the Cancer Care Ontario (CCO) Clinical Practice Guideline on active surveillance for the management of localized prostate cancer. Based on this guideline, we developed a deep learning model to classify prostate adenocarcinoma into indolent (applicable for active surveillance) and aggressive (necessary for definitive therapy) on core needle biopsy whole slide images (WSIs). In this study, we trained deep learning models using a combination of transfer, weakly supervised, and fully supervised learning approaches using a dataset of core needle biopsy WSIs (n=1300). We evaluated the models on a test set (n=645), achieving ROC-AUCs 0.846 (indolent) and 0.980 (aggressive). The results demonstrate the promising potential of deployment in a practical prostate adenocarcinoma histopathological diagnostic workflow system.

## 1 Introduction

According to the Global Cancer Statistics 2020, prostate cancer is the second most frequent cancer and the fifth leading cause of cancer death among men in 2020 Sung et al (2021). Prostate cancer is the most frequently diagnosed cancer in men in over one half (112 of 185) of the countries of the world Sung et al (2021). Therefore, it is necessary to define optimum therapeutic strategies for detection, treatment, and follow-up for prostate cancer patients Chen et al (2016). In recent year, pathologists perform the histopathological diagnosis of prostate cancer based on Gleason pattern quantities, tumor growth patterns, and clinical practice advancements (e.g., multiparametric magnetic resonance imaging (mpMRI) targeted biopsy and fusion ultrasound/magnetic resonance imaging biopsy) Van Leenders et al (2020). Standard active treatments for prostate cancer include hormone therapy, radiotherapy, and radical prostatectomy. However, to avoid the unnecessary side effects associated with overdiagnosis and over treatment, active surveillance is an important option for low-grade prostate cancer patients with reduced mortality risk Chen et al (2016); Morash et al (2015). As for the active surveillance, it consists in performing regular follow-ups of patients so as to be able to provide appropriate radical treatment for high-risk groups if necessary Morash et al (2015). The criteria for active surveillance are highly controversial Chen et al (2016); Morash et al (2015); Van Leenders et al (2020); Cyll et al (2022); Russell and Siddiqui (2022). According to the Cancer Care Ontario (CCO) Guideline and American Society of Clinical Oncology (ASCO) Clinical Practice Guideline, it is generally accepted that active surveillance is applied when a prostate cancer is determined by biopsy and Gleason pattern 4 components account for less than 10% of the total cancer volume Chen et al (2016). However, unfortunately, the inter-observer agreement for the Gleason score is not always high, and the inter-observer reproducibility (variability) of Gleason grading by general pathologists is often a problem Allsbrook Jr et al (2001); Oyama et al (2005); Ozkan et al (2016); Bulten et al (2022). Although International Society of Urological Pathology (ISUP) is making efforts to improve inter-observer agreement and equalize diagnostic quality for general pathologists by publishing consensus reviewing cases (https://isupweb.org/pib/), there are still cases that are not in agreement among pathologists in routine clinical practice.

In computational pathology, deep learning models have been widely applied in histopathological cancer classification on WSIs, cancer cell detection and segmentation, and the stratification of patient outcomes Yu et al (2016); Hou et al (2016); Madabhushi and Lee (2016); Litjens et al (2016); Kraus et al (2016); Korbar et al (2017); Luo et al (2017); Coudray et al (2018); Wei et al (2019); Gertych et al (2019); Bejnordi et al (2017); Saltz et al (2018); Campanella et al (2019); Iizuka et al (2020); Tsuneki et al (2022). Recently, it has been reported that an artificial intelligence (AI)-powered platform used as a clinical decision support tool was able to detect, grade, and quantify prostate cancer with high accuracy and efficiency and was associated with significant reductions in inter-observer variability Huang et al (2021); Bulten et al (2021). As for the global AI competition, the Prostate cANcer graDe Assessment (PANDA) challenge, a group of AI Gleason grading algorithms developed during a global competition generalized well to intercontinental and multinational cohorts with pathologist-level performance Bulten et al (2022). Other works Singhal et al (2022); Li et al (2021); Melo et al (2021); Otálora et al (2021); Silva-Rodríguez et al (2021); Marginean et al (2021); Nagpal et al (2019); Campanella et al (2019) have also looked into developing deep learning algorithms to classify prostate cancer Gleason scores based on histopathological images.

In this study, we investigated deep learning models to classify prostate adenocarcinoma in two classes based on the clinical responses: indolent (applicable for active surveillance) and aggressive (necessary for definitive therapy). To define the criteria of indolent and aggressive, we refered to CCO and ASCO guidelines Chen et al (2016) and set the cut-off value of 20% identified Gleason score 4 & 5 components in total prostate adenocarcinoma (Fig. 1) to reduce the possibility of inter-observer variability Sadimin et al (2016) as compared to the 10% cut-off value proposed by CCO and ASCO Chen et al (2016). To the best of our knowledge, this is the first study to establish a deep learning model to make an inference of the necessity for active surveillance on prostate core needle biopsy histopathology whole slide images (WSIs). We trained deep learning models using a combination of transfer learning, weakly, and fully supervised learning approaches and evaluated the trained models on core needle biopsy test set, achieving ROC-AUCs 0.846 (indolent) and 0.980 (aggressive). These findings suggest that it would be possible to not only detect adenocarcinoma on biopsy WSIs, but also to predict patients’ optimum clinical interventions (active surveillance or definitive therapy).

**Fig. 1:**
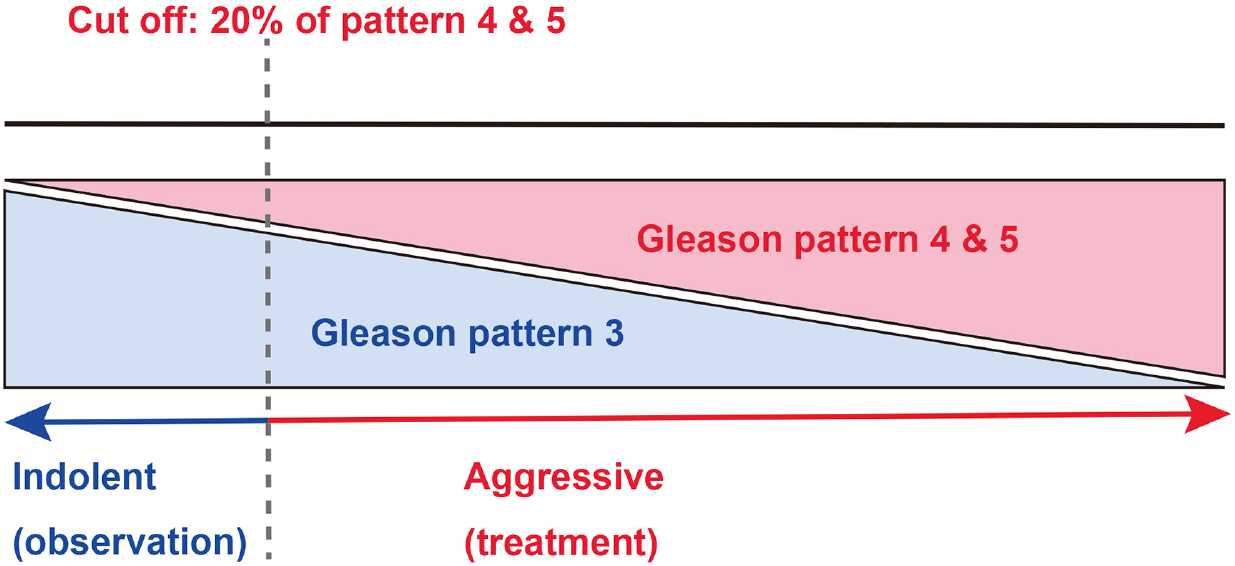
The schematic diagram of classification labels for prostate adenocarcinoma according to clinical treatment. If the whole slide image (WSI) with Gleason pattern 4 and 5 greater than or equal to 20% in the total area of prostate adenocarcinoma observed by pathologists, the WSI was classified as aggressive. On the other hand, the WSIs with Gleason pattern 4 and 5 less than 20% in the total area of prostate adenocarcinoma were classified as indolent.

## 2 Materials and methods

### 2.1 Clinical cases and pathological records

This is the retrospective study. A total of 2,285 H&E (hematoxylin & eosin) stained histopathological core needle biopsy specimen slides of human prostate adenocarcinoma and benign (non-neoplastic) lesions – 1,321 of adenocarcinoma and 964 of benign – were collected from the surgical pathology files of Kamachi Group Hospitals (Shinyukuhashi, Wajiro, and Shinkuki Hospitals) (Fukuoka, Japan) and Sapporo-Kosei General Hospital (Sapporo, Japan), after histopathological review of all specimens by surgical pathologists in each hospital. In Kamachi Group Hospitals, the histopathological specimens were selected randomly to reflect a real clinical settings as much as possible. In Sapporo-Kosei General Hospital, only adenocarcinoma specimens were provided. Prior to the experimental procedures, each WSI diagnosis was observed and verified by at least two senior pathologists. All WSIs were scanned at a magnification of x20 using the same Leica Aperio AT2 Digital Whole Slide Scanner (Leica Biosystems, Tokyo, Japan) and were saved as SVS file format with JPEG2000 compression.

### 2.2 Dataset

Table 1 shows breakdowns of the distribution of the specimens based on the following: all specimens, consensus specimens by two senior pathologists, training set, validation set, and test set of prostate core needle biopsy WSIs from Kamachi Group Hospitals and Sapporo-Kosei General Hospital. According to the Cancer Care Ontario Guideline Chen et al (2016) and American Society of Clinical Oncology (ASCO), patients with both low-volume (accounting for 10% total tumor) and intermediate-risk (Gleason score 3 + 4 = 7) prostate cancer may be offered active surveillance. At the same time, because of known inter-observer variability associated with the identification of minor Gleason pattern 4 components, prospective intradepartmental consultation with other pathologists should be considered for quality assurance Chen et al (2016). Therefore, in this study, considering clinical responses, we have set two classes for prostate adenocarcinoma: indolent and aggressive. Indolent suggests observation (active surveillance) and aggressive suggests definitive therapy.

**Table 1:** Distribution of cases in the different sets broken down by hospital and classification.

|  |  |  |  |
| --- | --- | --- | --- |
| All WSIs | Kamachi Group Hospitals | Sapporo-Kosei General Hospital | total |
| Adenocarcinoma | 718 | 603 | 1321 |
| Benign | 964 | 0 | 964 |
| total | 1682 | 603 | 2285 |
| Consensus | Kamachi Group Hospitals | Sapporo-Kosei General Hospital | total |
| Aggressive | 418 | 372 | 790 |
| Indolent | 81 | 140 | 221 |
| Benign | 964 | 0 | 964 |
| total | 1463 | 512 | 1975 |
| Training set | Kamachi Group Hospitals | Sapporo-Kosei General Hospital | total |
| Aggressive | 236 | 249 | 485 |
| Indolent | 24 | 87 | 111 |
| Benign | 704 | 0 | 704 |
| total | 964 | 336 | 1300 |
| Validation set | Kamachi Group Hospitals | Sapporo-Kosei General Hospital | total |
| Aggressive | 5 | 5 | 10 |
| Indolent | 5 | 5 | 10 |
| Benign | 10 | 0 | 10 |
| total | 20 | 10 | 30 |
| Test set | Kamachi Group Hospitals | Sapporo-Kosei General Hospital | total |
| Aggressive | 177 | 118 | 295 |
| Indolent | 52 | 48 | 100 |
| Benign | 250 | 0 | 250 |
| total | 479 | 166 | 645 |

In this study, we labelled (classified) prostate adenocarcinoma WSIs as follows. If the WSI has less than 20% of Gleason pattern 4 and Gleason pattern 5 components in total adenocarcinoma, it should be classified as indolent (Fig. 1). If the WSI has more than 20% of Gleason pattern 4 and Gleason pattern 5 components in total adenocarcinoma, it should be classified as aggressive (Fig. 1). We set the cut-off at 20% of total prostate adenocarcinoma on a WSI (Fig. 1) to reduce the possibility of interobserver variability as compared to 10% Chen et al (2016), because it has been widely reported that assessment of percentage Gleason pattern 4 in minute cancer foci has poor reproducibility among pathologists, especially for poorly formed glands Van Leenders et al (2020); McKenney et al (2011); Egevad et al (2011); Sadimin et al (2016); Zhou et al (2015); Harding-Jackson et al (2016).

In total we use indolent, aggressive, and benign as WSI labels for training the deep learning models at the WSI level. During the consensus review by two senior pathologists, 310 adenocarcinoma WSIs were excluded because of low concordance when classified into indolent or aggressive (Table 1). Training, validation, and test set were selected randomly from the consensus WSIs (Table 1).

### 2.3 Annotation

A senior pathologist, who performs routine histopathological diagnoses in general hospital, manually annotated 100 adenocarcinoma WSIs from the training set. The pathologist carried out annotations by free-hand drawing using an in-house online tool developed by customizing the open-source (OpenSeadragon) tool, which is a web-based viewer for zoomable images. On average, 10-15 lesions were annotated per WSI. The pathologists performed annotations based on the histopathological characteristics of Gleason pattern 3, 4, and 5. For example, well-formed glands with intraluminal crystalloids (Gleason pattern 3) (Fig. 2A), large irregular cribriform glands (Gleason pattern 4) (Fig. 2B), crowded fused glands (Gleason pattern 4) (Fig. 2C), poorly formed small-sized glands with some lumen-formation (Gleason pattern over 4) (Fig. 2D), ductal adenocarcinoma lined by columnar cells with elongated nuclei (Gleason pattern 4) (Fig. 2E), and infiltrating cords and single tumor cells without lumen formation (Gleason pattern 5) (Fig. 2F) were manually annotated. For training step, Gleason pattern 3 annotations were grouped as indolent and Gleason pattern 4 and 5 annotations as aggressive. The pathologist included cancer stroma which surrounds cancer cells in the annotation area. The average annotation time per WSI was about five minutes. All annotations performed by the pathologist were modified (if necessary), confirmed, and verified by a senior pathologist who performs routine histopathological diagnoses in general hospital.

**Fig. 2:**
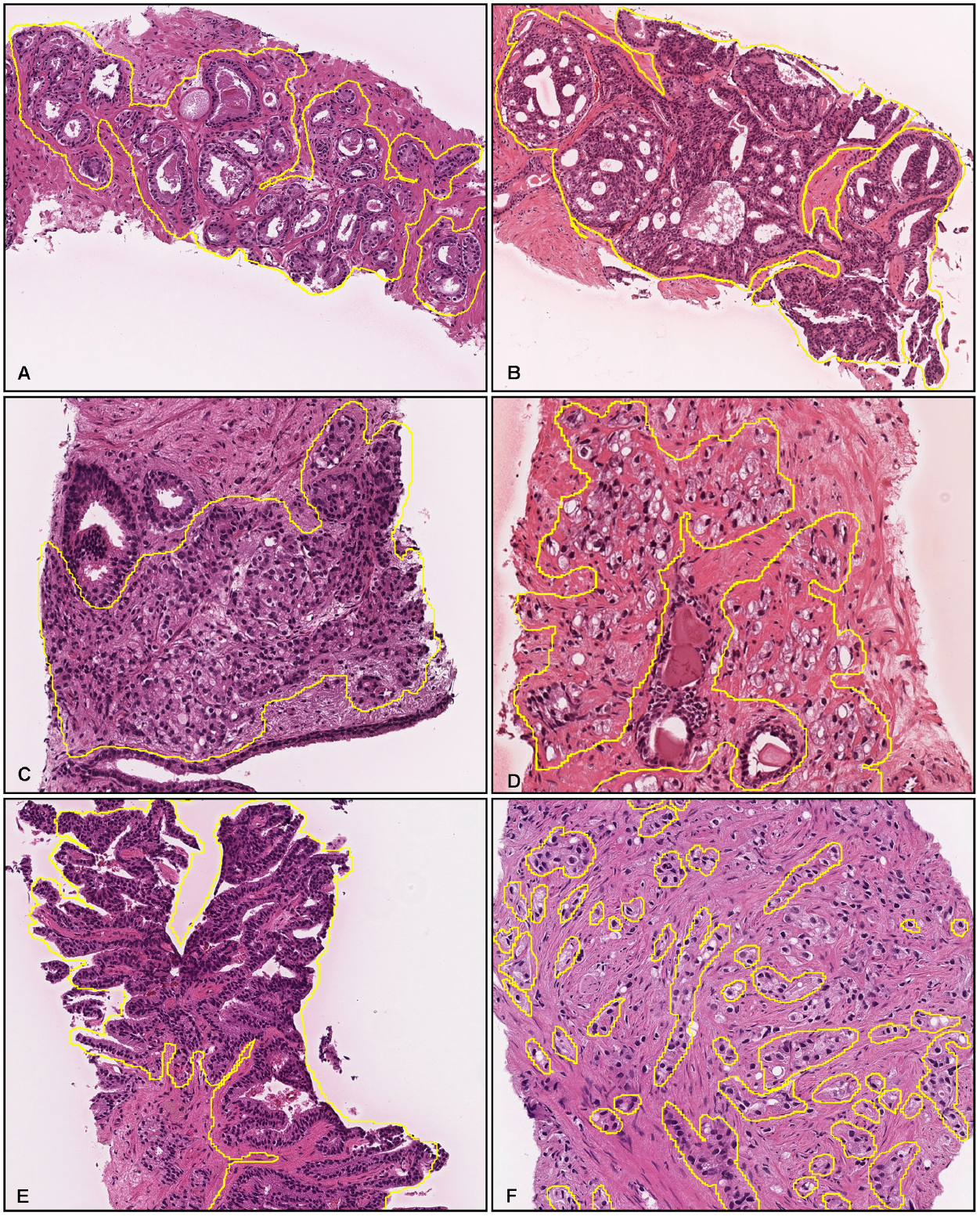
Representative images with manually-drawn annotations for Gleason pattern 3, 4, and 5 of adenocarcinoma. We performed annotations for wellformed glands with intraluminal crystalloids (Gleason pattern 3) (A), large irregular cribriform glands (Gleason pattern 4) (B), crowded fused glands (Gleason pattern 4) (C), poorly formed small-sized glands with some lumen-formation (Gleason pattern over 4) (D), ductal adenocarcinoma lined by columnar cells with elongated nuclei (Gleason pattern 4) (E), and infiltrating cords and single tumor cells without lumen formation (Gleason pattern 5) (F). We did not annotate areas where it was difficult to determine cytologically that the lesions were cancerous.

### 2.4 Deep learning models

We trained the models via transfer learning using the partial fine-tuning approachKanavati and Tsuneki (2021b). This is an efficient fine-tuning approach that consists of using the weights of an existing pre-trained model and only fine-tuning the affine parameters of the batch normalization layers and the final classification layer. For the model architecture, we used Efficient-NetB1Tan and Le (2019) starting with pre-trained weights on ImageNet. We used similar training methodology as Kanavati and Tsuneki (2021a); Tsuneki et al (2022). For clarity, we highlight the main parts below.

We performed tissue detection using Otsu’s thresholding method Otsu (1979) by excluding the white background. We then extracted tiles only from the tissue regions. During prediction, we extracted tiles from the entire tissue regions using a sliding window with a fixed-size stride. During training, we performed random balanced sampling of tiles, whereby we first randomly sampled three WSIs, one for each label. Then from each corresponding WSI, we randomly sampled an equal amount of tiles. For aggressive or indolent WSIs, we randomly sampled from the annotated tissue regions; for Benign, we randomly sampled from all the tissue regions.

After a few epochs, we switched to hard mining of tiles where we alternated between training and inference. During inference, the CNN was applied in a sliding window fashion on all of the tissue regions in the WSI, and we then selected the *k* tiles with the highest probability for being positive. This step effectively selects the tiles that are most likely to be false positives when the WSI is negative. The selected tiles were placed in a training subset, and once that subset contained *N* tiles, the training was run. We used *k* = 8, *N* = 256, and a batch size of 32.

For fully-supervised training, we performed the initial random sampling from annotated regions followed by the hard mining. We refer to this as FS+WS. For weakly-supervised training, we only performed the hard mining as it did not involve any annotations. We refer to this as WS.

To obtain a single prediction for the WSIs from the the tile predictions, we took the maximum probability from all of the tiles. We used the Adam optimizer Kingma and Ba (2014), with the binary cross-entropy as the loss function, with the following parameters: *beta*_1_ = 0.9, *beta*_2_ = 0.999, a batch size of 32, and a learning rate of 0.001 when fine-tuning. We used early stopping by tracking the performance of the model on a validation set, and training was stopped automatically when there was no further improvement on the validation loss for 10 epochs. We chose the model with the lowest validation loss as the final model.

### 2.5 Inter- and intra-rater reliability studies

To evaluate human pathologists’ inter-rater and intra-rater reliability, following WSIs were randomly selected from the test set: (i) 25 true negative WSIs (consensus classification by senior pathologists: Benign, deep learning model (TL-Colon poorly ADC (x20, 512) and FS+WS) WSI classification: Benign), (ii) 25 true-positive (indolent) WSIs (consensus: indolent, deep learning model: indolent), (iii) 25 false-positive WSIs (consensus: 13 indolent WSIs and 12 aggressive WSIs, deep learning model: 25 WSIs both indolent & aggressive double classes), (iv) 25 true-positive (aggressive) WSIs (consensus: aggressive, deep learning model: aggressive) (Table 4). A total of 100 WSIs were randomly shuffled and presented to volunteer pathologists using an in-house online tool developed by customizing the open-source (OpenSeadragon) tool, which is a web-based viewer for zoomable images. We performed the same intra-rater reliability study (Table 5) experiment twice with a one-month gap, randomising the order of WSIs each time. Volunteer pathologists recruited in this study consisted of 5 pathologists with less than 10 years experiences after becoming board certificated and 5 pathologists with more than 10 years experiences after becoming board certificated (total 10 pathologists) (Table 4).

### 2.6 Software and statistical analysis

The deep learning models were implemented and trained using TensorFlow Abadi et al (2015). AUCs were calculated in python using the scikit-learn package Pedregosa et al (2011) and plotted using matplotlib Hunter (2007). The 95% CIs of the AUCs were estimated using the bootstrap method Efron and Tibshirani (1994) with 1000 iterations.

The true positive rate (TPR) was computed as

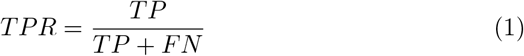

and the false positive rate (FPR) was computed as

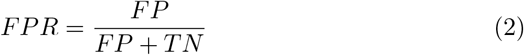

Where TP, FP, and TN represent true positive, false positive, and true negative, respectively. The ROC curve was computed by varying the probability threshold from 0.0 to 1.0 and computing both the TPR and FPR at the given threshold.

To assess the histopathological diagnostic concordance of pathologists, we performed S-score statistics, which is a measure and change-adjusted index for inter-rater reliability of categorical measurements between two or more raters Bennett et al (1954). To evaluate the intra-rater reliability for each pathologist, we performed the weighted kappa statistics Kundel and Polansky (2003); Swan et al (2022). We calculated the S-scores and kappa values using Microsoft Excel 2016 MSO (16.0.13029.20232) 64 bit. The scale for interpretation is as follows: *≤* 0.0, poor agreement; 0.01–0.20, slight agreement; 0.21–0.40, fair agreement; 0.41–0.60, moderate agreement; 0.61–0.80, substantial agreement; 0.81–1.00, almost perfect agreement (Tables 4, 5).

### 2.7 Availability of data and material

The datasets generated during and/or analysed during the current study are not publicly available due to specific institutional requirements governing privacy protection but are available from the corresponding author on reasonable request. The datasets that support the findings of this study are available from Kamachi Group Hospitals (Fukuoka, Japan) and Sapporo-Kosei General Hospital (Sapporo, Japan), but restrictions apply to the availability of these data, which were used under a data use agreement which was made according to the Ethical Guidelines for Medical and Health Research Involving Human Subjects as set by the Japanese Ministry of Health, Labour and Welfare (Tokyo, Japan), and so are not publicly available. However, the data are available from the authors upon reasonable request for private viewing and with permission from the corresponding medical institutions within the terms of the data use agreement and if compliant with the ethical and legal requirements as stipulated by the Japanese Ministry of Health, Labour and Welfare.

## 3 Results

### 3.1 High AUC performance of prostate core needle biopsy WSI evaluation of indolent and aggressive adenocarcinoma histopathology images

We trained deep learning models using two different training approaches: one was transfer learning (TL) and weakly supervised learning (WS) approach Kanavati et al (2020); Tsuneki et al (2022) (TL-Colon poorly ADC (x20, 512) and WS) and the other was TL and fully supervised (FS) pre-training followed by WS (FS + WS) approach Kanavati et al (2021) (TL-Colon poorly ADC (x20, 512) and FS + WS). Both approaches, the models were applied in a sliding window fashion with input tiles of 512×512 pixels, magnification at x20, and strides of 256. As for transfer learning, colon poorly differentiated adenocarcinoma classification model (Colon poorly ADC (x20, 512)) Tsuneki and Kanavati (2021) was selected as an initial weight due to its highest ROC-AUC (0.889, CI: 0.861 - 0.914) and lowest log-loss (0.415, CI: 0.378 - 0.457) (Table 2) on test set (Table 1). The other existing deep learning models (Table 2) we have used to compare ROC-AUC and log-loss performances were described previously: Stomach ADC, AD (x10, 512) Iizuka et al (2020); Stomach signet ring cell carcinoma (SRCC) (x10, 224) Kanavati et al (2021); Stomach poorly ADC (x20, 224) Kanavati and Tsuneki (2021a); Colon ADC, AD (x10, 512) Iizuka et al (2020); Pancreas EUS-FNA ADC (x10, 224) Naito et al (2021); Breast IDC, DCIS (x10, 224) Kanavati et al (2022). As for FS pre-training, we have used manually drawing annotations by pathologists 2.For test set (Table 1), we computed the ROC-AUC, log loss, accuracy, sensitivity, and specificity and summarized in Table 3 and Fig. 3.

**Table 2:** ROC-AUC and log loss results for aggressive classification on the core needle biopsy test set using existing adenocarcinoma classification models

| Existing deep learning models | ROC-AUC | Log loss |
| --- | --- | --- |
| Stomach ADC, AD (x10, 512) | 0.768 [0.734 - 0.808] | 1.443 [1.286 - 1.563] |
| Stomach SRCC (x10, 224) | 0.787 [0.747 - 0.823] | 0.858 [0.768 - 0.949] |
| Stomach poorly ADC (x20, 224) | 0.806 [0.771 - 0.840] | 0.542 [0.516 - 0.568] |
| Colon ADC, AD (x10, 512) | 0.568 [0.518 - 0.606] | 1.499 [1.371 - 1.665] |
| Colon poorly ADC (x20, 512) | 0.889 [0.861 - 0.914] | 0.415 [0.378 - 0.457] |
| Pancreas EUS-FNA ADC (x10, 224) | 0.739 [0.703 - 0.782] | 0.639 [0.596 - 0.677] |
| Breast IDC, DCIS (x10, 224) | 0.748 [0.705 - 0.784] | 1.450 [1.333 - 1.569] |

**Table 3:** ROC-AUC, log loss, accuracy, sensitivity, and specificity results for aggressive and indolent classification on the core needle biopsy test set using transfer learning (TL) and weakly supervised learning (WS) model (TL-Colon poorly ADC (x20, 512) and WS) and fully and weakly supervised learning model (TL-Colon poorly ADC (x20, 512) and FS+WS)

| TL-Colon poorly ADC (x20, 512) and WS |  |  |  |  |
| --- | --- | --- | --- | --- |
|  | aggressive |  | indolent |  |
| ROC-AUC | 0.970 | [0.957 - 0.981] | 0.851 | [0.819 - 0.885] |
| Log-Loss | 0.410 | [0.320 - 0.500] | 1.133 | [0.959 - 1.298] |
| Accuracy | 0.918 | [0.898 - 0.940] | 0.758 | [0.727 - 0.792] |
| Sensitivity | 0.885 | [0.846 - 0.920] | 0.870 | [0.798 - 0.933] |
| Specificity | 0.946 | [0.925 - 0.968] | 0.738 | [0.705 - 0.777] |

**Table 3:** ROC-AUC, log loss, accuracy, sensitivity, and specificity results for aggressive and indolent classification on the core needle biopsy test set using transfer learning (TL) and weakly supervised learning (WS) model (TL-Colon poorly ADC (x20, 512) and WS) and fully and weakly supervised ...
| TL-Colon poorly ADC (x20, 512) and FS+WS |  |  |  |  |
| --- | --- | --- | --- | --- |
|  | aggressive |  | indolent |  |
| ROC-AUC | 0.980 | [0.969 - 0.990] | 0.846 | [0.813 - 0.879] |
| Log-Loss | 0.213 | [0.160 - 0.260] | 2.273 | [2.012 - 2.475] |
| Accuracy | 0.935 | [0.918 - 0.957] | 0.736 | [0.707 - 0.772] |
| Sensitivity | 0.946 | [0.919 - 0.973] | 0.900 | [0.833 - 0.955] |
| Specificity | 0.926 | [0.899 - 0.955] | 0.706 | [0.673 - 0.750] |

**Fig. 3:**
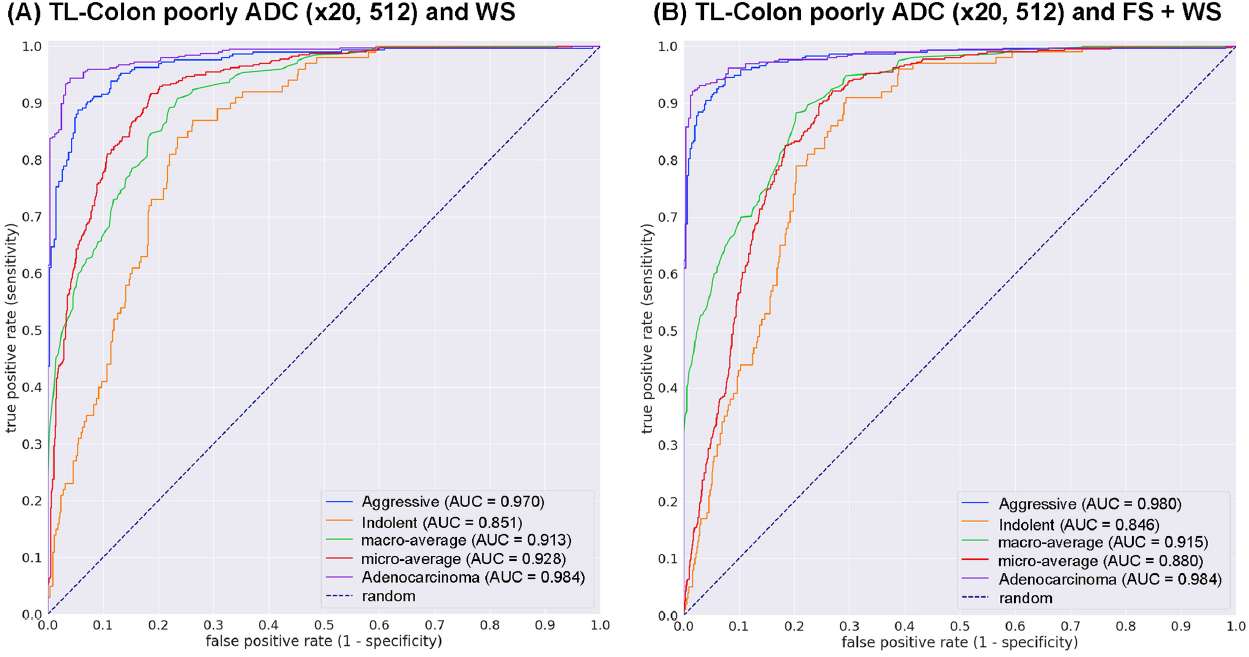
ROC curves with AUCs from two trained deep learning models on the test set. (A) transfer learning (TL) model from existing colon poorly differentiated adenocarcinoma (ADC) classification model with tile size 512 px and magnification at x20, following weakly supervised learning using training set with whole slide image (WSI) labeling. (B) TL model from existing colon ADC classification model with tile size 512 px and magnification at x20, following fully and weakly supervised learning using training set with annotation and WSI labeling.

As for WSI classification, the deep learning model for FS pre-training followed by WS approach (TL-Colon poorly ADC (x20, 512) and FS + WS) slightly improved ROC-AUC, accuracy, and sensitivity and decreased log-loss as compared to the model for WS approach (TL-Colon poorly ADC (x20, 512) and WS) in aggressive WSIs but not in indolent WSIs (Fig. 3 and Table 3). On the other hand, when compared with and without FS learning ([TL-Colon poorly ADC (x20, 512) and FS + WS] and [TL-Colon poorly ADC (x20, 512) and WS]) models for indolent and aggressive prediction at tile level in WSIs, FS pre-training followed by WS (FS + WS) approach robustly predicted indolent (Gleason pattern 3) (Fig. 4A, C, D, F) and aggressive (Gleason pattern 4 and 5) (Fig. 4M, O, P, R) patterns on heatmap images as compared to the WS approach (TL-Colon poorly ADC (x20, 512) and WS) (Fig. 4A, B, D, E, M, N, P, Q). Interestingly, the model (TL-Colon poorly ADC (x20, 512) and FS + WS) predicted indolent pattern (Gleason pattern 3) area precisely where pathologists did not mark ink-dots when they performed diagnosis (Fig. 4G, I, J, L), which was not predicted by the WS approach (TL-Colon poorly ADC (x20, 512) and WS) (Fig. 4G, H, J, K).

**Fig. 4:**
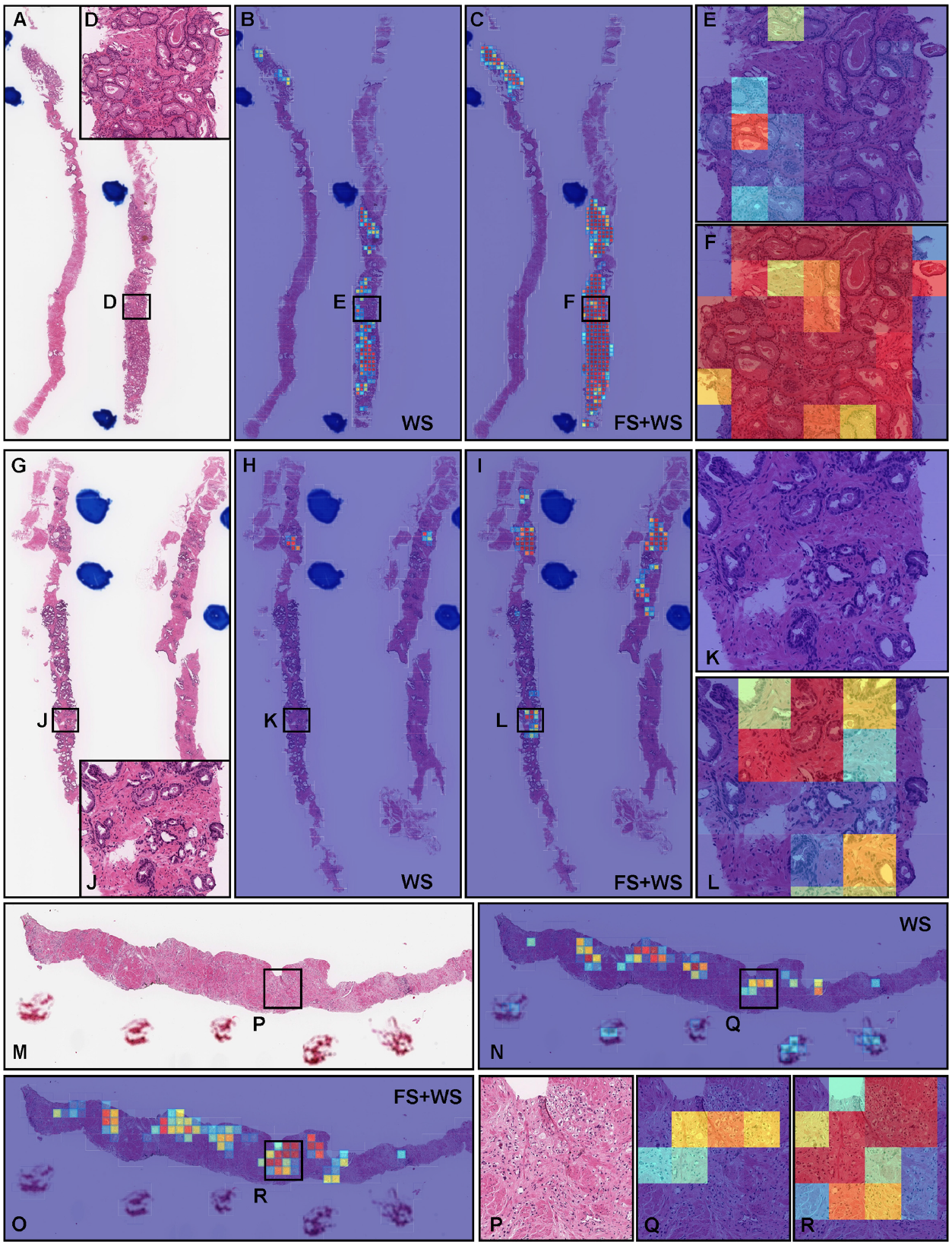
Comparison of indolent and aggressive prediction in core needle biopsy whole slide images (WSIs) of two trained deep learning models with and without fully supervised learning ([TL-Colon poorly ADC (x20, 512) and WS] and [TL-Colon poorly ADC (x20, 512) and FS+WS]). In (A), Gleason pattern 3 adenocarcinoma (D) was observed in all fragments. The heatmap images show indolent prediction outputs (B, C, E, F). As compared to the weakly supervised (WS) model (B, E), fully supervised (FS) and WS model predicted indolent morphology (Gleason pattern 3) more precisely (F) and indolent predicted area was almost same as pathologist’s marking with blue ink-dots. In (G), the pathologist had missed identifying Gleason pattern 3 adenocarcinoma in (J). WS model did not predict the presence of adenocarcinoma in the same area (K). FS+WS model predicted precisely indolent (Gleason pattern 3) area (L). In (M), infiltrating single cell adenocarcinoma (Gleason pattern 5) (P) was predicted correctly as aggressive (Q) by WS model. FS+WS model predicted infiltrating adenocarcinoma as aggressive more precisely (R). The heatmap uses the jet color map where blue indicates low probability and red indicates high probability.

Figures 5, 6, 7, 8 show representative WSIs of true-positive, true-negative, false-positive, and false-negative, respectively from using the model (TL-Colon poorly ADC (x20, 512) and FS + WS).

**Fig. 5:**
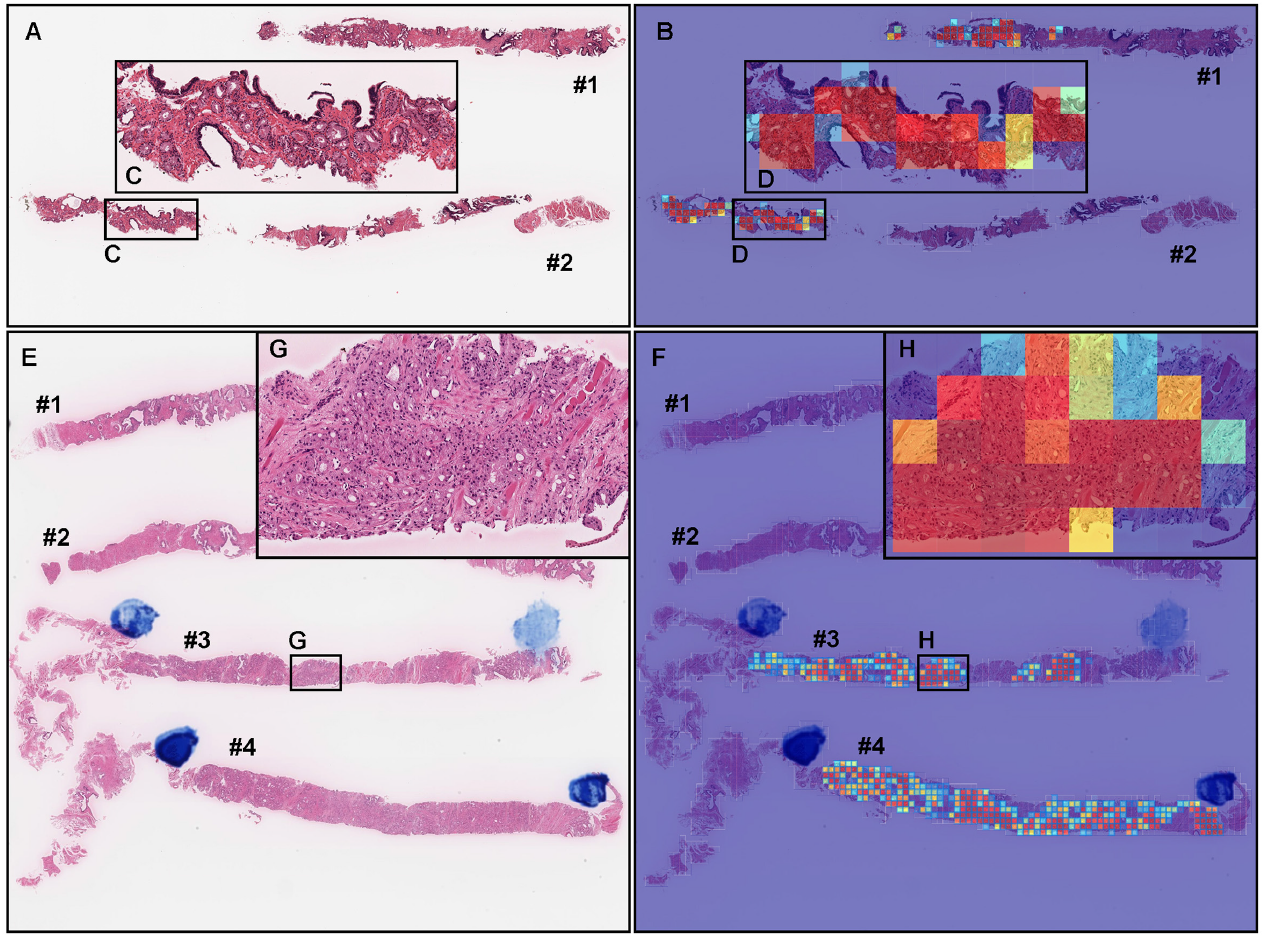
Two representative examples of indolent and aggressive true positive prediction outputs on whole slide images (WSIs) from core needle biopsy test set using the model (TL-Colon poorly ADC (x20, 512) and FS+WS). In the WSI of core needle biopsy specimen (A), histopathologically, adenocarcinoma corresponded with Gleason score 3+3 (C) infiltrated in both #1 and #2 fragments. The heatmap image (B) shows true positive indolent predictions (B, D) which correspond respectively to H&E histopathology (C, D). Histopathologically, in (E), #1 and #2 fragments were benign (non-neoplastic) lesions. Prostate adenocarcinoma which form small fused glands (G) corresponded with Gleason score 4+4 infiltrated in #3 and #4 fragments. The heatmap image (F) shows true positive aggressive predictions (F, H) which correspond respectively to H&E histopathology (E, G). The heatmap uses the jet color map where blue indicates low probability and red indicates high probability.

**Fig. 6:**
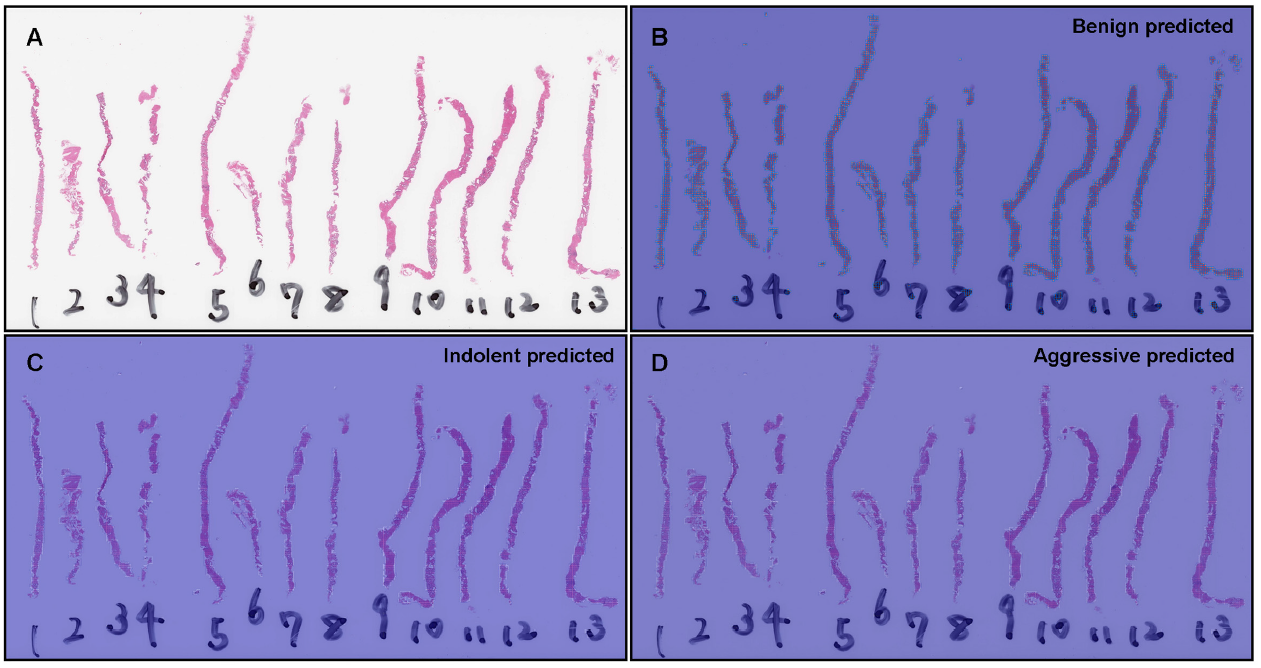
Representative true negative indolent and aggressive prediction outputs on a whole slide image (WSI) from core needle biopsy test set using the model (TL-Colon poorly ADC (x20, 512) and FS+WS). Histopathologically, in (A), all tissue fragments (#1-#13) were benign (non-neoplastic) lesions without any evidence of malignancy. The heatmap image (B) shows true positive predictions of benign and heatmap images, while (C) and (D) show true negative predictions of indolent and aggressive, respectively. The heatmap uses the jet color map where blue indicates low probability and red indicates high probability.

**Fig. 7:**
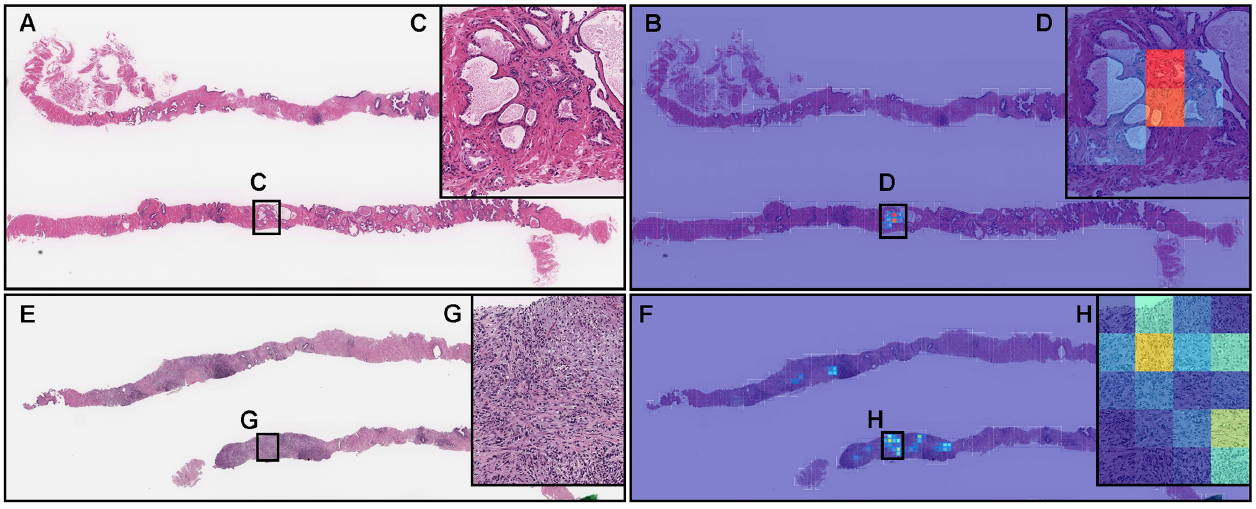
Two representative examples of indolent and aggressive false positive prediction outputs on whole slide image (WSIs) from core needle biopsy test set using the model (TL-Colon poorly ADC (x20, 512) and FS+WS). Histopathologically, (A, C) is a prostatic hyperplasia and (E, G) is a chronic prostatitis, both of which are benign (non-neoplastic) lesions. The heatmap image (B, D) exhibits false positive predictions of indolent (D) where the tissue consists of large and small dilated atrophic glands (C). The heatmap images (F, H) show false positive predictions of aggressive (H) where the tissue consists of severe infiltration of lymphocytes and histiocytes (G). The heatmap uses the jet color map where blue indicates low probability and red indicates high probability.

**Fig. 8:**
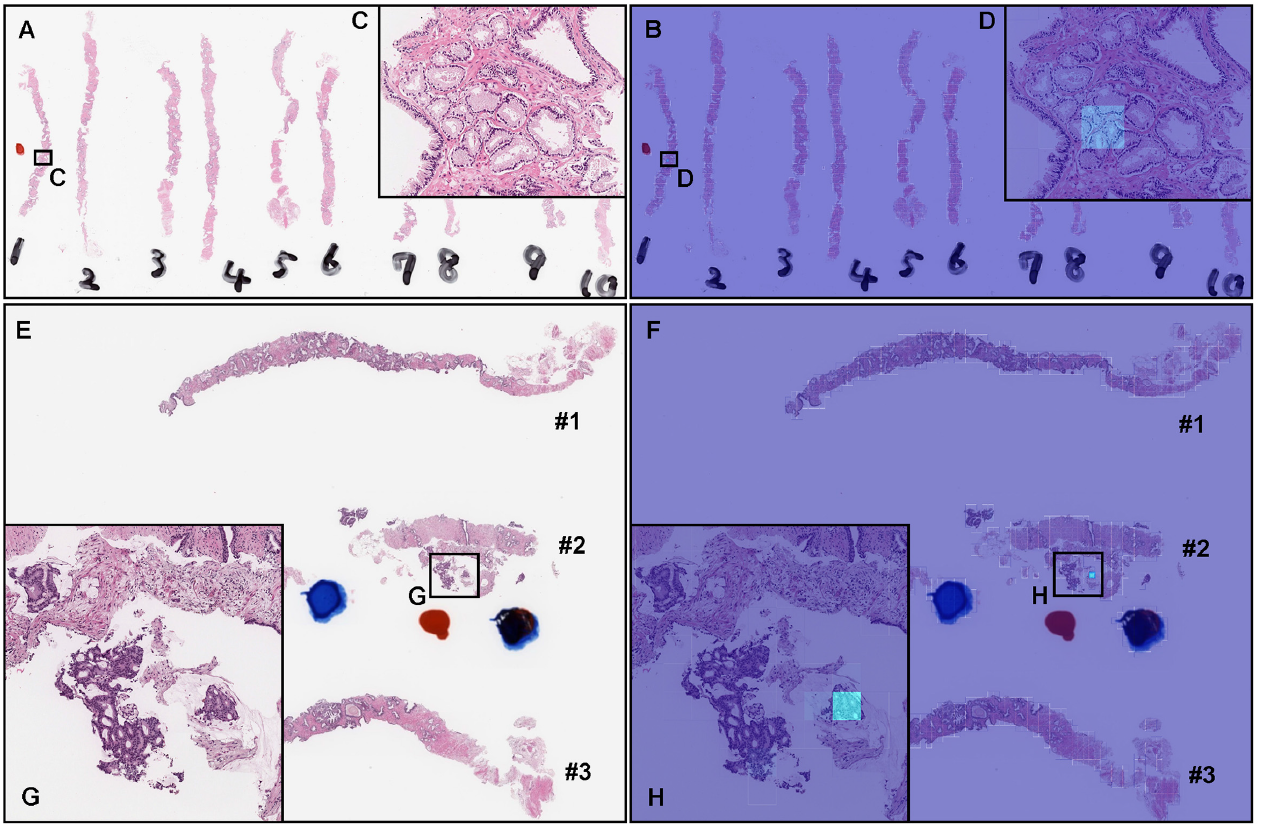
Two representative examples of indolent and aggressive false negative prediction output on whole slide images (WSIs) from core needle biopsy test set using the model (TL-Colon poorly ADC (x20, 512) and FS+WS). Histopathologically, in (A), infiltration of adenocarcinoma which exhibited Gleason pattern 3 was observed only in a limited area (C) of the #1 fragment where pathologist marked a red ink-dot on the glass slide. There was no evidence of malignancy in #2-#10 fragments (A). The heatmap image (B) show a true positive prediction of indolent on the Gleason pattern 3 adenocarcinoma (C) with very low probability (D). The prediction output at WSI level was benign (B). Histopathologically, in (E), there were a few fragmented adenocarcinoma foci with cribriform pattern which exhibited Gleason pattern 4 (G) in the #2 fragment. The heatmap image (F) show a true positive prediction of aggressive on a few adenocarcinoma (G) with very low probability (H). The prediction output at WSI level was benign (F). The heatmap uses the jet color map where blue indicates low probability and red indicates high probability.

### 3.2 True positive indolent and aggressive prediction of core needle biopsy WSIs

Our model (TL-Colon poorly ADC (x20, 512) and FS + WS) satisfactorily predicted indolent (Fig. 5A-D) and aggressive (Fig. 5E-H) patterns in core needle biopsy WSIs. According to the histopathological report and additional pathologists’ consensus reviewing, in both #1 and #2 tissue fragments (Fig. 5A), there are adenocarcinoma corresponded with Gleason pattern 3 (Gleason score = 3 + 3) (Fig. 5C), indicating indolent adenocarcinoma pattern and indolent WSI classification. The heatmap image (Fig. 5B, D) shows true positive indolent predictions in #1 and #2 fragments (Fig. 5B), where corresponded with H&E morphology (Fig. 5C, D). In (Fig. 5E), #1 and #2 fragments were benign (non-neoplastic) lesions and there are adenocarcinoma corresponded with Gleason pattern 4 (Gleason score = 4 + 4) (Fig. 5G), indicating aggressive adenocarcinoma pattern and aggressive WSI classification. The heatmap image (Fig. 5F, H) shows true positive aggressive predictions in #3 and #4 fragments (Fig. 5F), where corresponded with H&E morphology (Fig. 5G, H). False positive predictions were not observed in other benign tissue fragments (#1 and #2) (Fig. 5E, F).

### 3.3 True negative indolent and aggressive prediction of core needle biopsy WSIs

Our model (TL-Colon poorly ADC (x20, 512) and FS + WS) showed true negative predictions of indolent (Fig. 6A, C) and aggressive (Fig. 6A, D) patterns in core needle biopsy WSIs. In Fig. 6A, histopathologically, all tissue fragments (#1-#13) were benign (non-neoplastic) lesions. The heatmap image showed true positive prediction of benign (Fig. 6B), true negative predictions of indolent (Fig. 6C) and aggressive (Fig. 6D) patterns.

### 3.4 False positive indolent and aggressive prediction of core needle biopsy WSIs

According to the histopathological reports and additional pathologists’ reviewing, Fig. 7A is a prostatic hyperplasia and Fig. 7E is a chronic prostatitis, which are benign (non-neoplastic) lesions. Our model (TL-Colon poorly ADC (x20, 512) and FS + WS) showed false positive predictions of indolent (Fig. 7B) and aggressive (Fig. 7F) patterns, which caused indolent and aggressive WSI classification. indolent false positive tissue areas showed large and small dilated atrophic glands (Fig. 7C, D) and aggressive false positive tissue areas showed severe infiltration of lymphocytes and histiocytes (Fig. 7G, H), which could be the primary causes of false positives due to its morphological similarity in indolent pattern (Gleason pattern 3) and aggressive pattern (Gleason pattern 4 and 5).

### 3.5 False negative indolent and aggressive prediction of core needle biopsy WSIs

According to the histopathological reports and additional pathologists’ consensus reviewing, in Fig. 8A, infiltrating adenocarcinoma showed indolent pattern (Gleason pattern 3) in the limited area of fragment #1 (Fig. 8C). Fragment #2-#10 were benign (non-neoplastic) lesions. The heatmap image (Fig. 8B) showed a weakly indolent predicted tile (Fig. 8D) which was corresponded with Gleason pattern 3 histopathology (Fig. 8C). Therefore, the false negative WSI classification was provided. In Fig. 8E, a few fragmented adenocarcinoma foci with cribriform pattern which indicated aggressive pattern (Gleason pattern 4) (Fig. 8G) in a fragment (#2). The heatmap image (Fig. 8F) showed true positive prediction of a few adenocarcinoma with low probability (Fig. 8H). Therefore, the false negative WSI classification was provided. Both of these WSIs (Fig. 8A, E) consist of very low volume of adenocarcinoma, which could be the primary causes of false negatives.

### 3.6 Both indolent and aggressive prediction outputs of core needle biopsy WSIs

There were 114 out of 645 WSIs in the test set (Table 1) which were predicted as both indolent and aggressive by our model (TL-Colon poorly ADC (x20, 512) and FS + WS). After looking over these WSIs carefully, we found tendencies in these WSIs which consisted of mixture of Gleason pattern 3 and Gleason pattern 4 adenocarcinoma in degree of the borderline (cut-off 20%) between indolent and aggressive evaluation (Fig. 1). For example, histopathologically, small, indistinct, or fused glands (equivalent to Gleason pattern 4) adenocarcinoma was predominant (Fig. 9A, D, E). However, at the same time, Gleason pattern 3 adenocarcinoma was mixed in various degrees (Fig. 9A, D, E) in the area of Gleason pattern 4 adenocarcinoma infiltration. Importantly, in all 114 WSIs predicted as both indolent and aggressive predicted, the boundary between Gleason pattern 3 and Gleason pattern 4 adenocarcinoma was unclear and traditional which was confirmed retrospectively by senior pathologists. The heatmap images of indolent (Fig. 9B) and aggressive (Fig. 9C) revealed that to some extent, indolent (Gleason pattern 3) (Fig. 9F, H) and aggressive (Gleason pattern 4 and 5) (Fig. 9G, I) prediction outputs were overlapped. Therefore, the WSI prediction outputs (indolent or aggressive) were approximate values. In these WSIs, the WSI classification was selected larger value of indolent or aggressive. If we compute ROC-AUC and log-loss based on the criteria for acceptance of double label WSI classification outputs (meaning both indolent and aggressive prediction outputs), the scores are as follows: indolent ROC-AUC 0.956 [CI: 0.940-0.970], log-loss 0.969 [CI: 0.835-1.109]; aggressive ROC-AUC 0.980 [CI: 0.969-0.990], log-loss 0.213 [CI: 0.167-0.264].

**Fig. 9:**
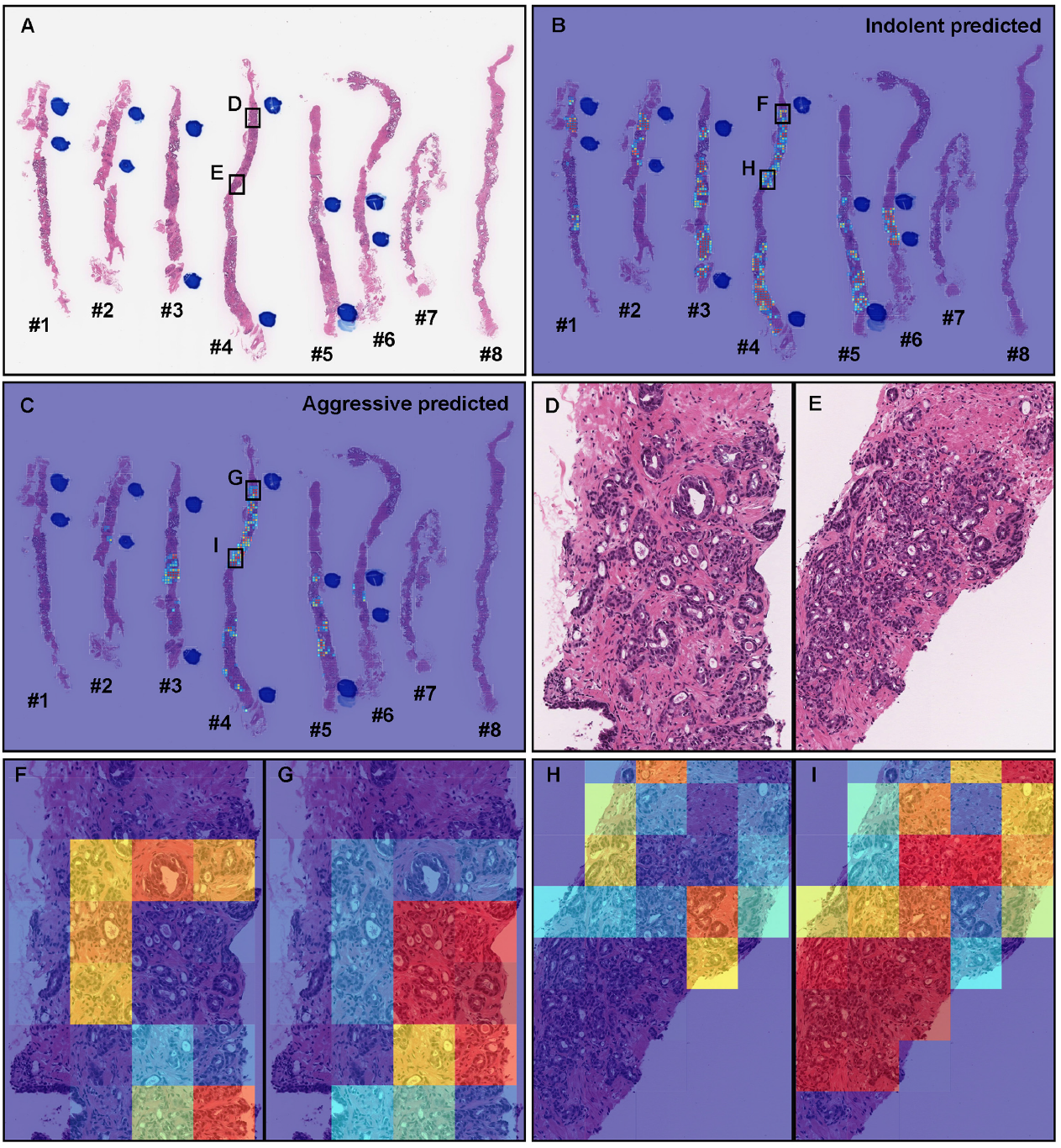
A representative example of a case that had both indolent and aggressive prediction outputs on a whole slide image (WSI) from the core needle biopsy test set using the model (TL-Colon poorly ADC (x20, 512) and FS+WS). In (A), small, indistinct, or fused glands (Gleason pattern 4) adenocarcinoma was predominant; however, Gleason pattern 3 adenocarcinoma is mixed in various degrees (D, E). The boundary between Gleason patterns 3 and 4 adenocarcinoma was unclear and transitional (D, E). The heatmap image (B) shows indolent prediction, and (C) shows aggressive prediction. In both (D) and (E) areas, indolent (F, H) and aggressive (G, I) prediction outputs were overlapped. The model (TL-Colon poorly ADC (x20, 512) and FS+WS) predicted the WSI (A) as both indolent and aggressive. The heatmap uses the jet color map where blue indicates low probability and red indicates high probability.

### 3.7 Inter and intra rater reliability study

To assess the inter-rater reliability of benign, indolent adenocarcinoma, and aggressive adenocarcinoma classification on WSIs, we have selected WSI based on our deep learning model (TL-Colon poorly ADC (x20, 512) and FS + WS) WSI prediction outputs and consensus classification by senior pathologists. As for true-negative cohort (25 WSIs; consensus: benign, AI predicted label: benign), S-scores in the range of 0.90-0.95, indicating “almost perfect agreement” (Table 4). As for the true-positive indolent cohort (25 WSIs; consensus: indolent, AI predicted label: indolent), S-scores in the range of 0.56-0.72, indicating “moderate to substantial agreement” (Table 4). As for the both indolent and aggressive predicted cohort (25 WSIs; consensus: 13 indolent and 12 aggressive, AI predicted label: indolent & aggressive), S-scores in the range of 0.10-0.28, indicating “slight to fair agreement” (Table 4). As for the true-positive aggressive cohort (25 WSIs; consensus: aggressive, AI predicted label: aggressive),S-scores in the range of 0.48-0.81, indicating “moderate to almost perfect agreement” (Table 4). The inter-rater reliability study was performed two times by randomizing a total 100 of identical WSIs with a one-month interval between 1st and 2nd studies. The S-scores in the 2nd study were slightly higher than 1st study and interpretations in the 2nd study were modestly improved than 1st study (Table 4). As for the aggressive classification, the S-scores in the pathologists more than 10 years experiences were higher than pathologists less than 10 years experiences (Table 4). Overall, WSIs which were predicted as both indolent & aggressive labels by our deep learning model (TL-Colon poorly ADC (x20, 512) and FS + WS) resulted very low S-scores in the range of 0.10-0.28, meaning poor inter-rater reliability (agreement) (Table 4) by pathologists regardless of experiences. As for the intra-rater reliability, all 10 pathologists achieved robust weighted kappa values in the range of 0.93-0.97, indicating “almost perfect agreement” (Table 5. Figure 10 shows a representative example WSI of poor evaluation (diagnostic) concordance among pathologists. As for the inter-rater reliability study, 5 pathologists evaluated as indolent and 5 pathologist as aggressive in this WSI (Fig. 10A). In Fig. 10A, there are wide variety of adenocarcinoma histopathologies. The heatmap images show both indolent (Fig. 10B) and aggressive (Fig. 10C) predictions by our deep learning model (TL-Colon poorly ADC (x20, 512) and FS + WS). In Fig. 10D, Gleason pattern 3 (indicating indolent) adenocarcinoma was predominant, which was predicted as indolent (Fig. 10E) not aggressive (Fig. 10F). In Fig. 10G and J, Gleason pattern 3 (indicating indolent) and Gleason pattern 4 (indicating aggressive) adenocarcinoma were mixed and it was hard to evaluate between two labels (indolent and aggressive), which were predicted as both indolent (Fig. 10H and K) and aggressive (Fig. 10I and L).

**Fig. 10:**
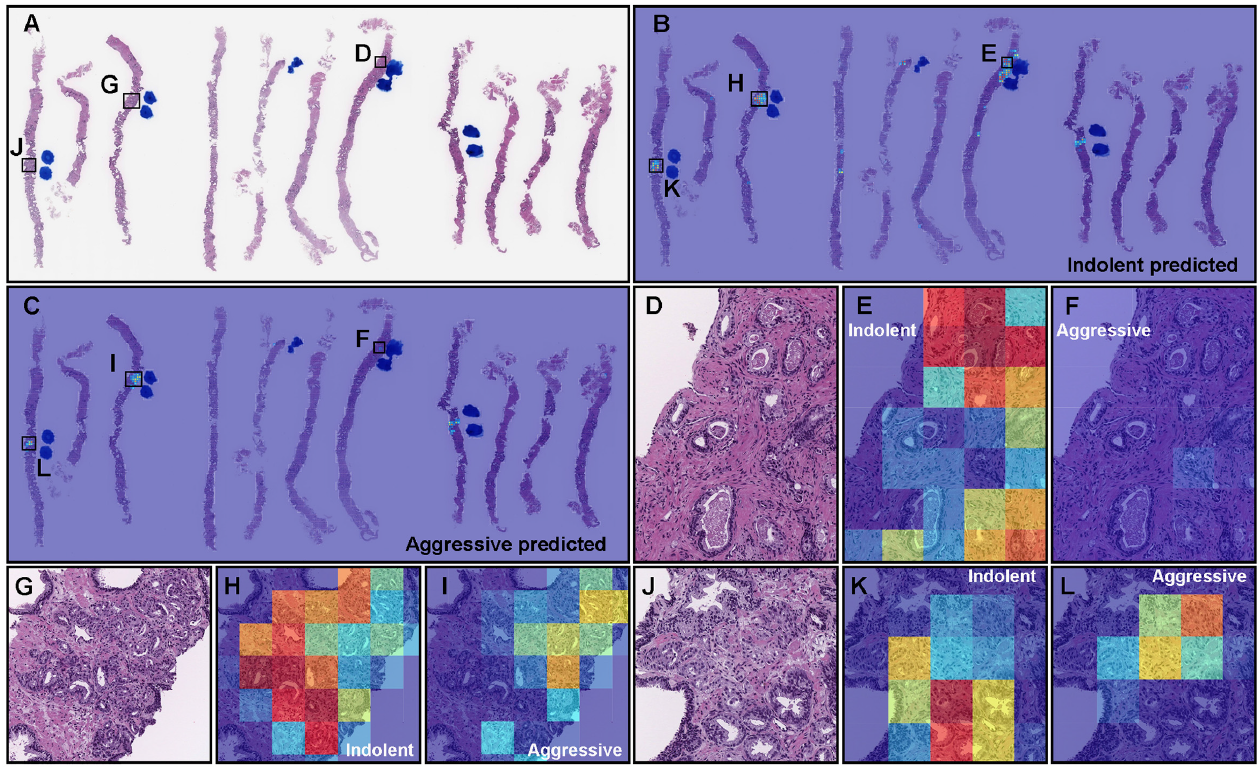
A representative example whole slide image (WSI) of poor evaluation (diagnostic) concordance among pathologists. Histopathologically, in (A), there were wide varieties of adenocarcinoma morphology. The heatmap image (B) shows indolent prediction and (C) shows aggressive prediction. In (D), Gleason pattern 3 adenocarcinoma was predominant, which was precisely predicted as indolent (E) but not as aggressive (F). In (G), Gleason pattern 3 and 4 adenocarcinoma were mixed, which were predicted as both indolent (H) and aggressive (I). In (J), the majority of adenocarcinoma was mixed Gleason pattern 3 and Gleason pattern 4, which were predicted as both indolent (K) and aggressive (L). The model (TL-Colon poorly ADC (x20, 512) and FS+WS) predicted the WSI (A) as both indolent and aggressive. The heatmap uses the jet color map where blue indicates low probability and red indicates high probability.

**Table 4:** Inter-rater reliability between pathologists using the S-score for two experiments on the same set conducted with a one month gap.

| 1st | Consensus | Predicted label | number of pathologists |  |  | interpretation |
| --- | --- | --- | --- | --- | --- | --- |
| | | | 10 (all) | 5 (< 10 yrs) | 5 (≥ 10 yrs) | |
|  | benign (25/25) | benign | 0.93 | 0.95 | 0.90 | almost perfect agreement |
|  | indolent (25/25) | indolent | 0.58 | 0.56 | 0.58 | moderate agreement |
|  | indolent (13/25), aggressive (12/25) | indolent & aggressive | 0.18 | 0.10 | 0.17 | slight agreement |
|  | indolent (13/25) | indolent & aggressive | 0.18 | 0.10 | 0.19 | slight agreement |
|  | aggressive (12/25) | indolent & aggressive | 0.18 | 0.10 | 0.15 | slight agreement |
|  | aggressive (25/25) | aggressive | 0.61 | 0.48 | 0.70 | moderate to substantial agreement |

| 2nd | Consensus | Predicted label | number of pathologists |  |  | interpretation |
| --- | --- | --- | --- | --- | --- | --- |
| | | | 10 (all) | 5 (< 10 yrs) | 5 (≥ 10 yrs) | |
|  | benign (25/25) | benign | 0.95 | 0.95 | 0.95 | almost perfect agreement |
|  | indolent (25/25) | indolent | 0.70 | 0.65 | 0.72 | substantial agreement |
|  | indolent (13/25), aggressive (12/25) | indolent & aggressive | 0.26 | 0.18 | 0.22 | slight to fair agreement |
|  | indolent (13/25) | indolent & aggressive | 0.28 | 0.19 | 0.26 | slight to fair agreement |
|  | aggressive (12/25) | indolent & aggressive | 0.23 | 0.18 | 0.18 | slight to fair agreement |
|  | aggressive (25/25) | aggressive | 0.75 | 0.68 | 0.81 | substantial to almost perfect agreement |

**Table 5:** Weighted kappa intra-rater scores for the 10 pathologists.

|  | weighted kappa |
| --- | --- |
| Pathologist-1 (< 10 yrs) | 0.97 |
| Pathologist-2 (< 10 yrs) | 0.94 |
| Pathologist-3 (< 10 yrs) | 0.95 |
| Pathologist-4 (< 10 yrs) | 0.93 |
| Pathologist-5 (< 10 yrs) | 0.95 |
| Pathologist-6 (≥ 10 yrs) | 0.97 |
| Pathologist-7 (≥ 10 yrs) | 0.94 |
| Pathologist-8 (≥ 10 yrs) | 0.94 |
| Pathologist-9 (≥ 10 yrs) | 0.95 |
| Pathologist-10 (≥ 10 yrs) | 0.96 |

## 4 Discussion

In this study, we trained deep learning models for the classification of indolent and aggressive prostate adenocarcinoma in core needle biopsy WSIs to make an inference for patients’ optimum clinical interventions (active surveillance or definitive therapy). We trained deep learning models using a combination of transfer learning Kanavati and Tsuneki (2021b); Tsuneki and Kanavati (2021); Tsuneki et al (2022), weakly supervised Kanavati et al (2020), and fully supervised Iizuka et al (2020); Kanavati et al (2021); Kanavati and Tsuneki (2021a) learning approaches. The evaluation results on the WSI level showed no significant differences between transfer learning and weakly supervised learning model (TL-Colon poorly ADC (x20, 512) and WS) and transfer learning, fully and weakly supervised learning model (TL-Colon poorly ADC (x20, 512) and FS+WS) (Table 3). However, the results at the tile level (visualised via heatmap images), the model (TL-Colon poorly ADC (x20, 512) and FS+WS) predicted both indolent (Gleason pattern 3) and aggressive (Gleason pattern 4 and 5) areas more precisely than weakly supervised learning model (TL-Colon poorly ADC (x20, 512) and WS) (Fig. 4. Therefore, we have selected the model (TL-Colon poorly ADC (x20, 512) and FS+WS) as the best model, which achieved ROC-AUCs at 0.846 (CI: 0.813 - 0.879) (indolent) and 0.980 (CI: 0.969 - 0.990) (aggressive) (Table 3). To the best of our knowledge, this is the first study to demonstrate the deep learning model to predict patients’ clinical interventions (active surveillance or definitive therapy) based on the histopathological WSIs. A previously reported deep learning model achieved ROC-AUC in the range of 0.855 (external test set) - 0.974 (internal test set) for the classification of benign and Gleason grade group 1-2 vs. Gleason grade group greater than or equal to 3 Bulten et al (2020). Our model (TL-Colon poorly ADC (x20, 512) and FS+WS) achieved better ROC-AUC performance in aggressive (0.980 (CI: 0.969 - 0.990)) (Table 3). These results suggest that the approach to predict patients’ clinical interventions could potentially achieve better deep learning model performance than the conventional Gleason score (grade) predicting approach. Our model (TL-Colon poorly ADC (x20, 512) and FS+WS) predicted indolent (Gleason pattern 3) and aggressive (Gleason pattern 4 and 5) lesions well after inspection of WSI heatmaps (Fig. 4, 5, 6). The model still had a few cases of false positive and false negative predictions (Fig. 7, 8). Our model (TL-Colon poorly ADC (x20, 512) and FS+WS) tends to show false positive predictions of indolent lesions where the tissues consist of atrophic glands and aggressive lesions where the tissues consist of severe inflammatory cell infiltration (Fig. 7). Our model tends to show false negative predictions of indolent and aggressive lesions where adenocarcinoma tissues were limited volumes (Fig. 8).

However, a major limitation (issue) in this study is that there was wide variability in inter-rater (observer) concordance among pathologists regardless of their years of experiences after becoming board certificated pathologists (Table 4), especially on the WSIs with both indolent (Gleason pattern 3) and aggressive (Gleason pattern 4 and 5) components mixed in various proportions (Table 4 and Fig. 10) Meliti et al (2017). On such WSIs which consisted of mixture of Gleason pattern 3 and Gleason pattern 4 adenocarcinoma in degree of the borderline (cut-off 20%) between indolent and aggressive evaluation (Fig. 1), our deep learning model (TL-Colon poorly ADC (x20, 512) and FS+WS) tends to predict both indolent and aggressive WSI outputs (17.7% of total WSIs in the test set) as well as pathologists (Fig. 9, 10). Indeed, there were a certain number of WSIs with Gleason pattern 4 or Gleason pattern 5 component around 20% of total adenocarcinoma in the test set, which were the major cause of poor concordance among pathologists and deep learning model WSI prediction outputs with both indolent and aggressive (Fig. 10). It has been reported that with less than 10% involvement of the core, it was more difficult to assess in smaller foci, with only moderate agreement Sadimin et al (2016). Given that in a small focus only a few glands of a given pattern can markedly affect the percent Gleason pattern 4, consideration should be given to not recording percent Gleason pattern 4 in small foci of Gleason score 7 tumors on core needle biopsy Sadimin et al (2016). This issue is inevitable when classifying WSIs based on percentages of adenocarcinoma components (Gleason pattern 3, 4, 5). Moreover, there were a certain number of WSIs in which there was a marked discrepancy among pathologists as to whether the prostate adenocarcinoma was classified as Gleason pattern 3 or Gleason pattern 4 (Fig. 10). Practically, the histopathological segregation of Gleason pattern 3 and Gleason pattern 4 is often problematic Egevad et al (2011); Meliti et al (2017). Currently, according to the diagnostic criteria of Gleason pattern 4 adenocarcinoma on core needle biopsy, poorly formed glands immediately adjacent to other well-formed glands regardless of their number and small foci of less than or equal to 5 poorly formed glands regardless of their location should be graded as Gleason pattern 3 Zhou et al (2015), which would be one of the primary cause of both indolent and aggressive prediction outputs. Moreover, in this study, instead of assigning an indolent or aggressive label to each core needle biopsy specimen, we considered all specimens on a WSI together as a single specimen Therefore, it was possible to be poor inter-observer concordance among pathologists if total histopathological area was too large (e.g., six or eight core specimens in a single WSI) to evaluate. However, it can be possible to resolve the issue by specimen preparation with one core needle biopsy specimen per glass slide (WSI) for biopsy specimens assuming the deep learning model prediction. Interestingly, when we compute the model (TL-Colon poorly ADC (x20, 512) and FS+WS) performance based on the criteria for acceptance of double label WSI classification outputs (both indolent and aggressive), indolent ROC-AUC were increased (0.956 [CI: 0.940-0.970]) and log-loss was decreased (0.969 [CI: 0.835-1.109]) as compared to Table 3. The other limitation in this study is that limited generalization of the deep learning model (TL-Colon poorly ADC (x20, 512) and FS+WS) because training and test set were provided by the same supplier hospitals (Kamachi Group Hospitals and Sapporo-Kosei General Hospital). Therefore, in the next step, to verify the versatility of the model (TL-Colon poorly ADC (x20, 512) and FS+WS), we need to perform verification study using enough number of WSIs from diverse range of hospitals.

The main advantage of our deep learning model (TL-Colon poorly ADC (x20, 512) and FS+WS) is that the model can predict patients’ optimum clinical interventions (active surveillance: indolent or definitive therapy: aggressive) on core needle biopsy WSIs. For most patients with low-risk (Gleason score less than or equal to 6) prostate cancer, active surveillance is the recommended disease management strategy Chen et al (2016). At the same time, select patients with low-volume, intermediate-risk prostate cancer (indolent in this study) can be offered active surveillance Chen et al (2016). In routine histopathological diagnosis for prostate cancer in core needle biopsy specimens, pathologists have to report Gleason scores for each core for risk assessment by using microscope which would be fatigue and laborious works. Moreover, it is revealed that there are significant inter-rater variability among pathologists in diagnosis of prostate cancer Sadimin et al (2016); Ozkan et al (2016); Meliti et al (2017). By using our deep learning model as an initial screening, pathologists can check WSIs with heatmap image highlighting indolent (Gleason pattern 3) and aggressive (Gleason pattern 4 and 5) adenocarcinoma and WSI prediction outputs (benign, indolent, and aggressive), which would be a great benefit for general pathologists to make diagnoses.

## 5 Acknowledgements

We are grateful for the support provided by Dr. Shigeo Nakano at Kamachi Group Hospitals (Fukuoka, Japan). We thank pathologists who have been engaged in reviewing cases and clinicopathological discussion for this study. This study is based on results obtained from a project, JPNP14012, subsidized by the New Energy and Industrial Technology Development Organization (NEDO).

## 6 Compliance with Ethical Standards

The experimental protocol was approved by the ethical board of Kamachi Group Hospitals (No. 173) and Sapporo-Kosei General Hospital (No. 597). All research activities complied with all relevant ethical regulations and were performed in accordance with relevant guidelines and regulations in the all hospitals mentioned above. Informed consent to use histopathological samples and pathological diagnostic reports for research purposes had previously been obtained from all patients prior to the surgical procedures at all hospitals, and the opportunity for refusal to participate in research had been guaranteed by an opt-out manner.

## 7 Funding

This study is based on results obtained from a project, JPNP14012, subsidized by the New Energy and Industrial Technology Development Organization (NEDO).

## 8 Conflict of Interest

F.K. and M.T. are employees of Medmain Inc. All authors declare no competing interests.

## 9 Contributions

M.T., M.A., S.I., and F.K. designed the studies. M.T. and F.K. performed experiments and analyzed the data; M.T. and F.K. performed the computational studies; M.A. and S.I. performed the histopathological diagnoses and reviewed the cases; M.T., M.A., and F.K. wrote the manuscript; M.T. supervised the project. All authors reviewed and approved the final manuscript.

## Notes

### Competing Interest Statement

M.T. and F.K. are employees of Medmain Inc. All authors declare no competing interests.

